# A knowledge graph approach to discovering drug combination therapies across the phenome

**DOI:** 10.1101/2025.05.13.25327531

**Authors:** Jianfeng Ke, Tingjian Ge, Rachel D. Melamed

## Abstract

Combining two clinically approved drugs has potential to improve treatment for common disease. But, with many thousands of combinations possible, clinically testing all pairs of drugs, with all common diseases, is not feasible. Here, we propose DRACO, a new machine learning method for discovering therapeutic drug combinations by leveraging knowledge about which health conditions each drug has been tested for. Our graph neural network model is able to generalize from a curated set of drug combinations to discover new combination therapies. We showcase our approach’s power to answer the question: given a drug and a health condition, what second drug would create an effective combination? Of our top three predicted candidate combinations, 74% have been previously reported. In the harder task of distinguishing the small number of reported combinations from millions of possible combination-condition triplets, our model ranks 94% of unseen triplets at the highest 0.1%. We expect DRACO to be a useful tool for proposing new therapies across thousands of diseases.

## Introduction

Drug combinations offer significant advantages in treating complex diseases[1], but discovering new combinations through clinical trials is both costly and time-consuming[2]. Therefore, it is crucial to leverage computational methods to pre-select promising drug combinations with a high likelihood of success before proceeding to clinical experimentation. Over the past decade, numerous models have been developed to predict drug combinations[2], [3]. While most focus on classifying or predicting combination synergy for specific cancers or cancer cell lines, only a few aim to evaluate the efficacy of drug combinations across different diseases. We propose that graph-based machine learning models can address this challenge due to their ability to integrate knowledge from diverse domains[2]. Recent studies have used graph learning to incorporate diverse biological knowledge—such as protein–protein interactions, drug indications, and side effects—and predict new drug effects[4], [5]. However, to our knowledge, no existing model transfers the ability to predict single drug indications to predict effective combinations of two drugs. In this study, we apply transfer learning by leveraging pre-trained drug and condition embeddings from the Drug Repurposing Knowledge Graph (DRKG)[6], which integrates data from 6 established databases, alongside a graph of drugs and the diseases they are tested on in clinical trials. We fine-tune the embeddings and train our model on the Continuous Drug Combination Database (CDCDB)[7]. Our model encompasses 1,943 drugs and 2,955 conditions, and outputs a probability score quantifying the predicted efficacy of any given drug pair for a specific condition.

## Methods Data curation

We curate 95,682 clinical trials from ClinicalTrials.gov (NIH), encompassing 1,943 distinct drugs and 2,955 conditions, where each trial represents the application of a single drug to a single condition. Of these, 1,831 drugs and 2,482 conditions have corresponding pre-trained embeddings available in the Drug Repurposing Knowledge Graph (DRKG; https://github.com/gnn4dr/DRKG). To construct the training dataset, which consists of samples pairing two drugs with one condition, we select 1,043 trials from ClinicalTrials.gov and 30,825 entries from the CDCDB (https://icc.ise.bgu.ac.il/medical_ai/CDCDB/), filtering both sources to include only combinations involving the investigated drugs and conditions. This results in a final dataset of 31,852 samples used for model development.

### Embeddings training and fine-tuning

We construct a bipartite graph where drugs and conditions are represented as nodes, and edges indicate that a drug has been tested for a condition in clinical trials. These drug–condition pairs are divided into two groups: the first group includes pairs where both the drug and the condition have pre-trained embeddings from the DRKG, and the second group includes all remaining pairs. To train embeddings for nodes lacking DRKG representations and to adapt the pre-trained ones to our study, we adopt a two-stage link prediction strategy. In the first stage, we randomly initialize the embeddings of nodes without DRKG representations and update only these, keeping the DRKG embeddings fixed. The goal of the first stage is to allow the randomly initialized embeddings to learn from the pre-trained DRKG embeddings by aligning their representation quality. Training continues until the median dot product of drug–condition pairs in the second group matches that of the first group (Figure 1A). Once this alignment is achieved, we proceed to the second stage to jointly fine-tune all embeddings in a task-specific manner. Both DRKG and newly learned embeddings are updated until the median dot products of the two groups converge again (Figure 1B). This two-stage link prediction strategy enables the integration of new node representations while refining the pre-trained embeddings.

**Figure 1.**
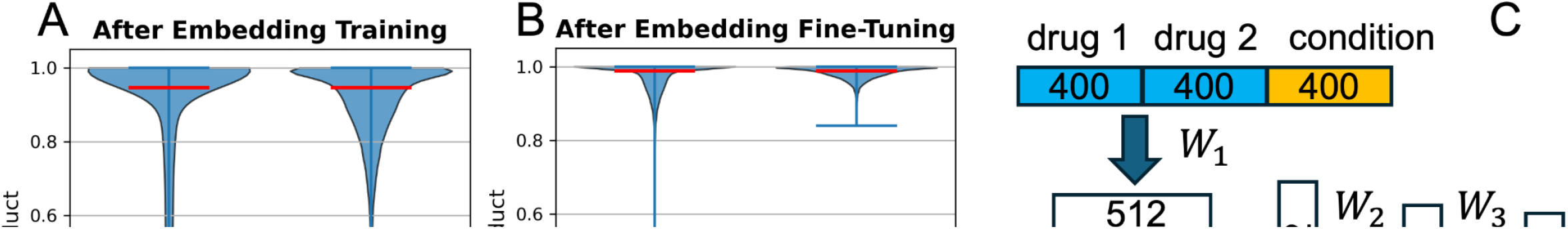
**(A)** Violin plot of the dot products between the embeddings of drug-condition pairs in two groups after embedding training. Group 1 includes pairs where both the drug and condition have DRKG embeddings; Group 2 includes all remaining pairs. **(B)** Violin plot of the dot products between drug-condition pair embeddings in the same two groups after embedding fine-tuning. **(C)** Schematic diagram of the model architecture.

We use a learning rate of 0.001 for embedding initialization in stage one and 0.0001 for fine-tuning in stage two. In both stages, newly trained and fine-tuned embeddings are clipped each epoch to the range defined by the minimum and maximum values of the DRKG embeddings. Gradients are clipped to a maximum norm of 1.0. For the 95,682 drug–condition pairs, we use a batch size of 8,192 in both stages, and optimize using binary cross-entropy loss (BCELoss).

### Model architecture and training

To predict the efficacy of a drug pair for a given condition, our model first represents all drugs and conditions using a shared embedding layer. Each input sample consists of a triplet: two drugs and one condition. To model the interaction among two drugs and the condition, we concatenate the fine-tuned embeddings of Drug 1, Drug 2, and the condition, and pass the resulting vector through a feedforward neural network, which captures the directional effect of the drug pair on the condition. To account for potential asymmetry in drug ordering, we compute the effect twice—once in the order (Drug 1, Drug 2, Condition), and once in the reversed order (Drug 2, Drug 1, Condition)—and sum the two outputs to produce an aggregated interaction representation. This aggregated effect vector is then fed into a second feedforward MLP, which outputs a final prediction score between 0 and 1, representing the model’s estimated efficacy of the drug combination for the given condition. Both MLP blocks include ReLU activations and dropout for regularization. The schematic diagram of the model architecture with the dimensionality of each layer is shown in Figure 1C. We randomly split the samples into 80% for training and 20% for testing. The initial learning rate is set to 0.001 and decays by a factor of 10 every 100 epochs. The model is trained for a total of 500 epochs, with a dropout rate of 0.5 and a batch size of 2,048.

### Negative sampling strategy

The CDCDB dataset used for training does not include explicit negative samples, that is, drug combinations known to be ineffective for specific conditions. Therefore, we generate negative samples during training. Observed drug–drug–condition triplets are extremely sparse relative to the full combinatorial space: with approximately 2,000 drugs and 3,000 conditions, the number of possible triplets approaches six billion. If negative samples are drawn uniformly at random, two forms of bias can arise. In the first case, a drug pair known to be effective for one condition is paired with a different condition. In the second case, a drug–condition pair from a known positive triplet is combined with another drug. In both cases, the resulting triplets contain components that frequently appear in the positive training data. As a result, the model may overfit to these recurring sub-patterns and misinterpret partially correct triplets—a drug combination targeting a condition—as valid. In addition, due to the extreme imbalance between the number of observed positive triplets and the space of potential negatives, we generate substantially more negative samples than positives to help the model distinguish effective combinations from ineffective ones during training. In our study, we use a negative-to-positive sampling ratio of 100:1. To reduce sampling bias and improve generalization, we regenerate negative samples at the start of each training epoch.

To address overfitting to partially correct triplets, we implement a negative sampling strategy that generates informative and structured negative examples. For the first case, where the model may overfit on a drug pair known to be effective, we construct negative samples by pairing the drug combination with a condition previously associated with one of the drugs in clinical trials. If no such condition exists, a condition is randomly selected from the global pool of conditions not yet linked to either drug. These samples are generated at a negative-to-positive ratio of 20:1. For the second case, where the model may overfit on a triplet that includes a drug–condition pair from a known positive sample combined with another drug, we construct negative samples by fixing the drug–condition pair and introducing a second drug. This additional drug is selected from those previously associated with the same condition in clinical trials. If no such drug exists, one is randomly drawn from the global pool of drugs not yet linked to that condition. These samples are generated at a negative-to-positive ratio of 80:1. To ensure unbiased model evaluation, we generate negative samples for the test set prior to training and exclude all such triplets—both test positives and their corresponding negatives—from the training process. The 20:1 and 80:1 sampling ratios for the two cases, respectively, were selected based on model performance on the test set, as they provided the best balance between generalization and ranking accuracy.

## Results

### Model Overview

We propose DRACO (Drug combinAtion for COnditions), a multi-layer perceptron (MLP)-based model designed to predict the efficacy of drug pairs for conditions. DRACO is, to our knowledge, the first model that transfers predictive knowledge from single-drug indications to drug combination prediction. It leverages pre-trained drug and condition embeddings from the Drug Repurposing Knowledge Graph (DRKG), while also learning embeddings for drugs and conditions not covered by DRKG and fine-tuning all embeddings for the combination prediction task (see Methods). Given two drugs and a condition, DRACO takes their embeddings as input and outputs a probability representing the predicted efficacy of the drug combination to the condition. By grounding the model in biologically meaningful embeddings and transferring knowledge from single-drug usage, DRACO enables robust prediction even in settings with limited labeled data.

### Model Evaluation on the testing set

To evaluate model performance after training, we first assess whether the model assigns higher predicted scores to known (existing) triplets of two drugs in a combination, plus the condition they treat. These scores are compared to negative unknown (unreported) triplets generated by fixing one drug and the condition.

Specifically, for each positive triplet, we fix one drug and the condition, and evaluate the rank of the positive triplet—containing the second drug—among all possible alternative, unreported drug combinations for that condition. Figure 2A presents bar plots of the rank distributions for both training and testing samples of triplets. Among more than 1,900 candidate drugs, the model ranks the true second drug within the top 3 for 94% of training samples and 74% of testing samples. These results demonstrate that, given a query health condition and a single drug, the model allows us to query for what second drug would form an effective combination therapy.

**Figure 2.**
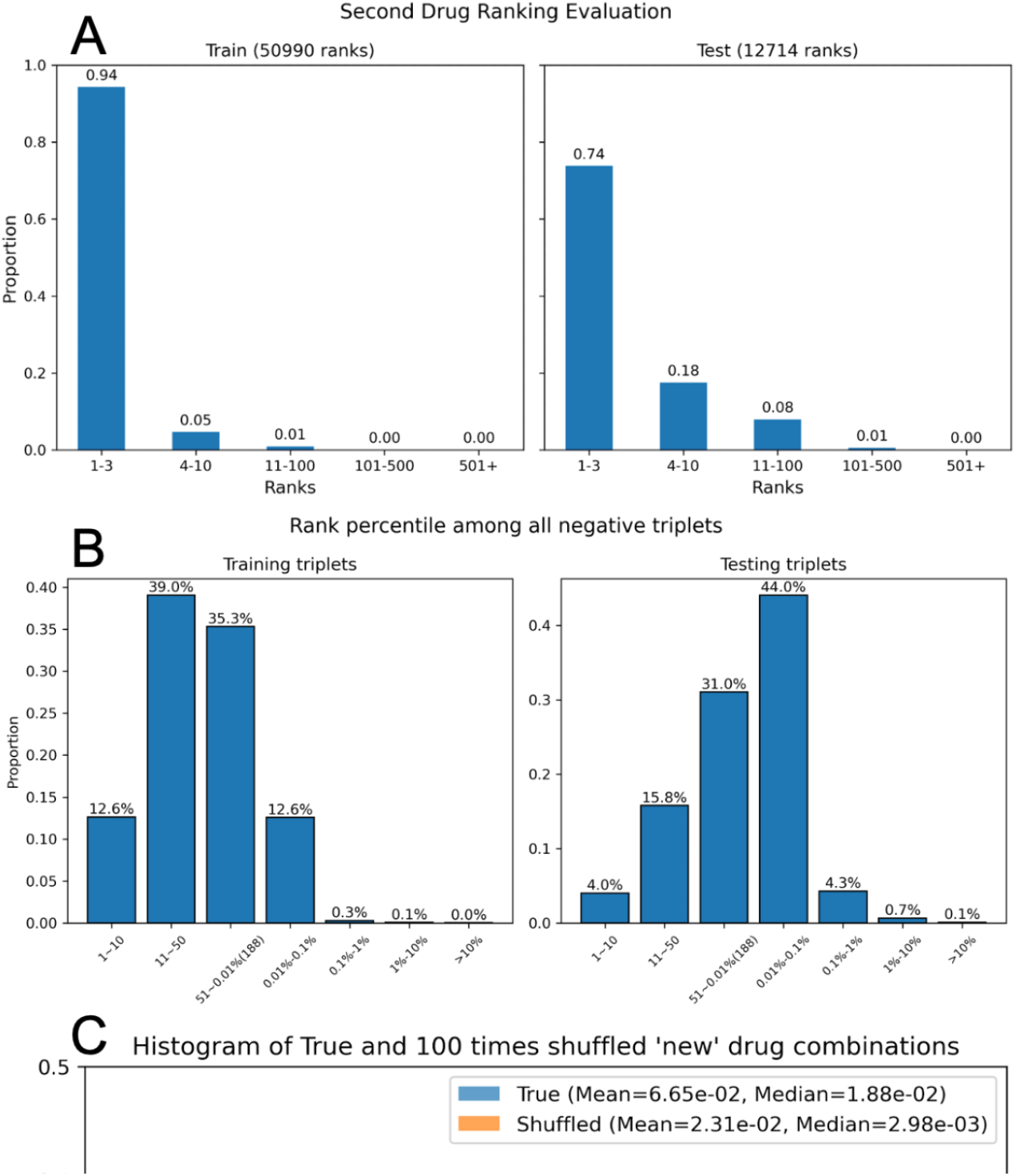
**(A)** Ranking of the second drug given a fixed first drug and condition. **(B)** Ranking of known drug combinations among all possible drug pairs for a fixed condition. **(C)** Distribution of potential scores for true vs. randomly shuffled drug combinations.

Next, we evaluate whether the model assigns higher predicted scores to known drug combinations compared to all possible alternative, unreported combinations for a given condition. With nearly 2,000 drugs, this results in approximately 2 million possible drug combinations for each condition, making this a stringent test. As shown in Figure 2B, over 94% of testing samples fall within the rank of top 0.1%, demonstrating the model’s strong capacity to prioritize true drug combinations over a large candidate space, with consistent performance maintained on the held-out testing set.

### Model Evaluation on unseen drug combination

To further assess the generalization ability of our model, we evaluate it on 2,552 drug combinations from CDCDB, none of which were present in our dataset, applied to conditions that were also not included in our study. A robust model is expected to assign higher scores to annotated drug combinations that were unseen during model development. To quantify this, we define the potential score of a drug combination as the highest predicted score across all conditions. We compare the potential scores of the 2,552 true drug combinations with those of randomly shuffled combinations. Specifically, we fix the first drug and randomly shuffle the second drug across the 2,552 pairs, repeating this process 100 times. To ensure a fair comparison, we exclude any shuffled combinations that were seen during training. Figure 2C shows the distribution of potential scores for both the true and shuffled combinations. Notably, 21% of the true drug combinations have a potential score between 0.1 and 1, compared to only 5.6% of the shuffled combinations within the same range. These results indicate that our model can successfully identify meaningful drug combinations.

## Discussion

DRACO combines knowledge of the medical conditions each drug has been tested on, alongside a relatively small curated list of drug combinations, in order to propose new drug combination therapies. Our graph model also allows us to incorporate pre-trained embedding representations of each drug and condition, which integrate multiple external information such as connections of drugs to genes and pathways. As our drug combination data set contains only positive reports of combination effects, a key design choice is the creation of negative example drug combinations that force the model training to generalize well to prediction of new drug combinations. Using a series of increasingly stringent measures of DRACO’s performance, we show that 1) it is highly successful at predicting which second drug could effectively combine with a first drug to treat a disease; 2) distinguishes previously reported drug combinations from those never reported. Previous work in this area has included efforts to find adverse effects of drug combinations, or to find combinations that could synergistically treat a particular patient’s tumor. To our knowledge, our approach is the first effort to comprehensively predict effective combinations across thousands of health conditions and drugs. Our approach can be used both to enhance the performance of currently used drugs for a disease, and to suggest new therapeutic approaches.

## Data Availability

All results are based on publicly available data.

## Notes

### Competing Interest Statement

The authors have declared no competing interest.

### Funding Statement

This work was supported by the National Institute of General Medicine Sciences (NIGMS R35 GM151001-01) to RDM.

